# Comparison of 2D Speckle Tracking Echocardiography and Cardiac MRI Feature Tracking for Bi-Atrial Strain Assessment in Elite Athletes

**DOI:** 10.1101/2025.08.19.25334025

**Authors:** J.J.N. Daems, M.A. van Diepen, J.C. van Hattum, S.M. Verwijs, B.J. Bouma, H.A.C.M. de Bruin – Bon, M.H. Moen, N.H. Prakken, A. van Randen, S.M. Boekholdt, M. Groenink, F.W. Asselbergs, H.T. Jorstad

## Abstract

**Background:** Data on Feature-tracking Cardiac Magnetic Resonance imaging (FT-CMR) in athletes is limited, and direct comparison of Speckle Tracking Transthoracic Echocardiography (STE) and FT-CMR-derived bi-atrial strain are lacking.

**Aim:** To compare STE and FT-CMR longitudinal bi-atrial strain in a well-defined cohort of elite athletes and to determine the level of agreement between these modalities.

**Methods:** Agreement between FT-CMR and STE derived bi-atrial strain was assessed in elite athletes from the Evaluation of Lifetime participation in Intensive Top-level sports and Exercise (ELITE) cohort (>16 years; exercise >10h/week; (inter)national or Olympic level). Absolute agreement was evaluated using a two-way mixed-effects model (ICC(3,1)).

**Results:** We included 194 elite athletes (51% women; mean age 30 ±7 years). The interval between TTE and CMR was <24h in 93.3% of athletes. There was poor-to-moderate absolute agreement in two-way-mixed-effects model with single measure reliability in left atrial reservoir-(0.34 (95% CI: 0.21 – 0.46, *p* < .001), conduit-(0.39 (95% CI: 0.26 – 0.50, p < .001), and contractile strain (0.48 (95% CI: 0.36 – 0.58, p < .001). There was no agreement in right atrial reservoir-(0.02, 95% CI: -0.12 – 0.16, *p* = .4) and conduit strain (0.09, 95% CI: -0.07 – 0.21, *p* = .17. There was poor agreement in RA contractile strain (0.218, 95% CI 0.080 – 0.48, *p* = 0.001).

**Conclusion:** There are important modality-dependent differences in bi-atrial strain in elite athletes. The poor-to-moderate agreement between longitudinal STE and FT-CMR left atrial strain and a lack of agreement in right atrial strain highlights the need to interpret observed values in the context of modality-specific elite athlete reference ranges. Clinicians should avoid direct cross-modality comparison in elite athlete populations.

## INTRODUCTION

Atrial enlargement is a feature of the athlete’s heart (1–7). However, atrial dilation is also strongly associated with numerous cardiovascular diseases, including primary atrial diseases (8–11). Alongside volume, atrial strain is emerging as an important tool to assess atrial function, and to distinguishing exercise-induced cardiac remodelling (EICR) from atrial pathology in athletes (1,3,5,8–11). Atrial strain quantifies the percentage change in the atrial myocardium during the three distinct phases of atrial function: reservoir, conduit, and contractile (12,13). Both transthoracic echocardiography (TTE) 2D speckle tracking (STE) and cardiac magnetic resonance imaging (CMR) feature tracking can be used to assess atrial strain. Most studies investigating atrial strain in athletes have primarily utilized STE due to its widespread availability and cost-effectiveness but STE is inherently limited by its dependence on intercostal windows, image quality, and operator expertise (14). Conversely, CMR is considered the reference standard for evaluating cardiac function and is increasingly used in the cardiovascular screening and clinical assessment of athletes (15,16). Moreover, feature-tracking CMR (FT-CMR) enables strain analysis with superior spatial resolution, greater reproducibility, and reduced reliance on acoustic windows (14).

Despite these advantages, FT-CMR studies investigating atrial strain in athletes are lacking, and most studies investigating STE-atrial strain in athletes have predominantly been performed in male endurance athletes (4,7,8). The agreement between STE and FT-CMR-derived atrial strain also remains poorly defined, as these assessments are both modality and software dependent (14,17). Reported agreement ranges from moderate to excellent, with conflicting findings across studies. (14,17). In addition, data on FT-CMR atrial strain in athletes is limited, and direct comparisons between STE- and FT-CMR-derived atrial strain in this population with substantial atrial remodelling are lacking. Establishing the level of agreement between these modalities in athletes is essential for reliable clinical interpretation and research applicability.

Therefore, we aimed to investigate and compare STE and FT-CMR bi-atrial strain in a well-defined cohort of elite athletes, and to investigate the level of agreement between these imaging modalities.

## METHODS

### Study design and population

We performed a cross-sectional, paired comparison of FT-CMR and STE derived bi-atrial strain in elite athletes to assess overall agreement, bias and limits of agreement.

### Population

We included elite athletes from the Evaluation of Lifetime participation in Intensive Top-level sports and Exercise (ELITE) cohort. The rationale and methods of the ELITE study have been described elsewhere (18). In short, ELITE collects data from standardized cardiovascular screenings of elite athletes who are older than 16 years of age, exercise >10h/week and play at the highest national, international, Olympic or Paralympic level). Screening includes, but is not limited to, TTE and CMR. Elite athletes were classified according to the European Society of Cardiology (ESC) sports classification (2). All consecutive athletes who underwent both a CMR and TTE until March 2024 were eligible for inclusion.

### TTE

Athletes underwent echocardiography with a GE E9 or E95 machine (GE Healthcare, Horten, Norway) with a 1.6-MHz to 3.2-MHz transducer (GE Healthcare, Milwaukee, WI) in left lateral decubitus position by licensed cardiac sonographers. ECG-gated 2D-greyscale, colour- and tissue Doppler recordings were made according to the guidelines of the European Association of Cardiovascular Imaging (EACVI) (19). Left atrial volumes were calculated following the area-length method in apical four-(4CH) and two-chamber (2CH) views. The right atrial volume was obtained in an apical four-chamber view, calculated using a single plane area-length.

### Speckle-tracking

Two operators experienced with STE bi-atrial strain computation analysed the TTE dataset in EchoPAC (version 6.0, GE Healthcare software) using the apical 2CH and 4CH with the onset of the QRS as zero reference point. Each study was analyzed by one operator only; analyses were not performed in duplicate. LA strain was determined from biplane view by automatic tracing of the endocardial borders of the LA, extrapolating across the pulmonary veins and/or LA appendage orifices (20). RA strain was derived from the 4CH by terracing the endocardial border of the RA lateral wall, RA roof, RA septal wall and ending at the opposite tricuspid annulus (20). Endocardial tracing was manually adjusted when necessary. Longitudinal bi-atrial strain was subdivided in reservoir-, conduit- and contractile strain [**Figure 1**].

**FIGURE 1.**
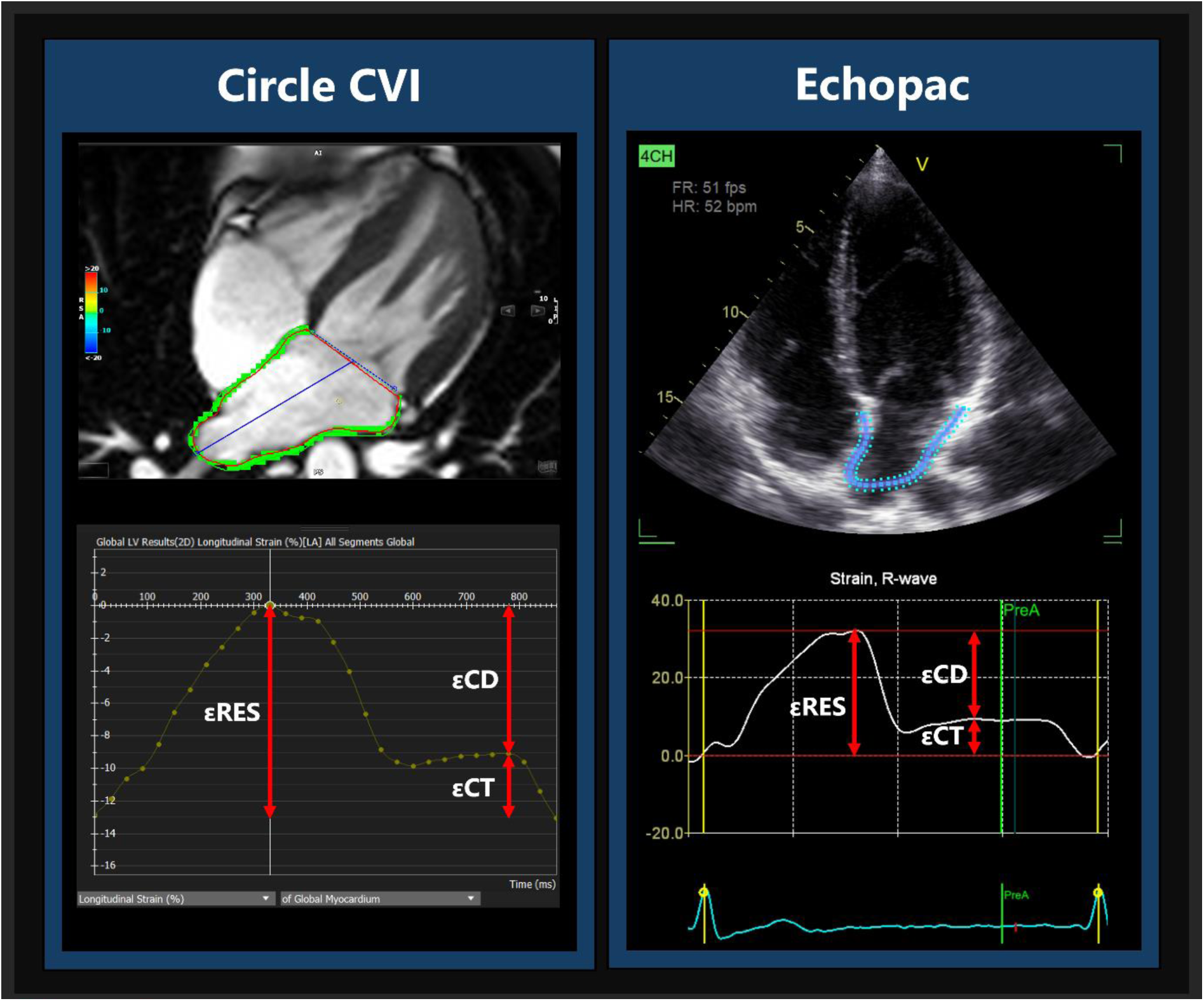
FT-CMR and STE Atrial Reservoir, Conduit and Contractile strain – εRes = Reservoir; εCD = Conduit ; εCT = Contractile

### CMR acquisition protocol

All CMRs were performed on a Siemens Avanto Fit 1.5 Tesla MRI scanner (Siemens, Erlangen, Germany) with dedicated 18-channel anterior and 32-channel posterior RF receive coils and standard ECG equipment (Siemens) for cardiac measurements and electrocardiography gating. The imaging protocol consists of standard cardiac localizers followed by balanced Steady-State Free Precession (bSSFP) CINE pulse sequences in long-axis views. Subsequently, a native T1 mapping modified look locker inversion recovery (MOLLI) sequence with 5(3)3 acquisition scheme was performed. Thereafter a gadolinium contrast bolus of 0.2 ml/Kg (Dotarem®, Guerbet, Roissy, France) was administered, followed by an imaging stack of contiguous short-axis bSSFP CINE slices, covering both ventricles from apex to base. Eight minutes after contrast administration, LGE imaging is performed. Fifteen minutes after contrast administration a post-contrast T1-mappingsequence with 4(1)3(1)2 MOLLI acquisition scheme was performed. See **[Supplementary Material 1**] for detailed MRI sequence parameters of the whole acquisition protocol.

### Feature tracking

A single operator experienced with FT-CMR derived atrial strain computation analysed the CMR dataset with the onset of the QRS as zero reference point. The operator was blinded to the results of STE. All CMR derived strain measurements were derived by a single observer. Longitudinal bi-atrial strain was quantified with the Feature Tracking module in Circle Cardiovascular Imaging (CVI version 5.17.1; build 3504; Calgary, Canada). Endo- and epicardial contours of the LA were traced in the end-systolic and diastolic phase in 2-chamber and 4-chamber cine images. RA monoplane derived longitudinal strain was derived by tracing the endo- and epicardial contours in the 4-chamber cine images. RA biplane derived longitudinal strain was derived by also tracing the endo- and epicardial contours in the RVOT outflow tract cine-images. When the atrial myocardium upon visual inspection was not traced properly by the automatic tracking algorithm, contours were adjusted manually. Longitudinal atrial bi-strain was subdivided in reservoir-, conduit- and contractile strain.

### Metrics of interest

Our primary metric of interest was agreement in longitudinal STE and FT-CMR bi-atrial strain, expressed as percentage of change of the atrial myocardium subdivided in atrial reservoir, conduit and contractile phase. Secondary metrics of interest comprised functional and dimensional atrial and ventricular features.

### Statistics

Continuous variables are presented as mean ± SD or median (IQR), and categorical variables as counts and percentages. Between-group differences were tested using independent t-tests or Mann-Whitney U tests for comparisons between women and men. Paired comparisons, such as TTE and CMR parameters measured in the same individuals, were analysed using paired t-tests or Wilcoxon signed-rank tests for continuous variables. Categorical variables were compared using χ² tests or Fisher’s Exact tests, as appropriate. Additionally, the rates of abnormal strain detected by TTE and CMR were compared with McNemar’s test for paired categorical data.

To assess the agreement between STE and FT-CMR strain we used correlation- and Bland-Altmann analysis. Spearman correlation was used to assess the association between STE and FT-CMR bi-atrial strain. Bland-Altmann analysis was used to assess bias, defined as the mean difference between TTE and CMR, and limits of agreement (LoA: bias ± 1.96 × SD). Absolute agreement was evaluated using a two-way mixed-effects model (ICC(3,1)), appropriate for single-measurements and fixed observers. Spearman’s Rho and ICC values were interpreted as weak/poor (<0.50), moderate (0.50–0.75), good (0.75–0.90), or excellent (>0.90). Differences between STE and FT-CMR strain were calculated for all components and subgroups. Their association with atrial size was assessed using correlation analysis.

### Comparison to Reference Values

Longitudinal left atrial strain was considered outside the reference range if values exceeded the respective population’s 95% confidence interval. For speckle-tracking echocardiography (STE), reference values were derived from a meta-analysis of 40 studies including 2542 healthy non-athletic individuals (LA reservoir 95% CI: 38.0% – 40.8%; LA conduit 95% CI: 20.7%–25.2%; LA contractile 95% CI: 16.0%–19.0%) (21). For feature-tracking cardiac magnetic resonance (FT-CMR), thresholds were based on a normative study of 112 healthy volunteers (22). While sex-specific normative values were available for FT-CMR, the reference ranges for STE atrial strain are only reported as pooled values. To maintain consistency in comparison we applied the pooled reference values for both STE (LA reservoir 95% CI: 38.0% – 40.8%; LA conduit 95% CI: 20.7%–25.2%; LA contractile 95% CI: 16.0%–19.0%) and FT-CMR (LA reservoir 95% CI: 37.9% – 40.9%; LA conduit 95% CI: 23.6% –26.7%; LA contractile 95% CI: 13.2% – 14.8%). McNemar’s test was used to compare the prevalence of values exceeding the reference range between STE and FT-CMR. At present, there are no robustly validated reference values for RA strain using either STE or FT-CMR. McNemar’s test was used to compare the prevalence of values exceeding the reference range between STE and FT-CMR. Currently there are no robustly validated reference values for RA strain for both TTE and CMR available.

### TTE inter-rater agreement

We calculated an intraclass correlation coefficient (ICC) two-way random-effects model with absolute agreement (ICC[2,1]) to evaluate single-measure based inter-rater agreement in STE strain. Data from 20 randomly selected athletes were analyzed, each assessed by both raters.

## RESULTS

### Athlete characteristics

We included a total of 194 elite athletes, whereof 51% women, with a mean age of 30 ±7 years. According to the ESC sports classifications, athletes competed primarily in 42% endurance-, 42% mixed-, 9.3% power-, and 6.7% skill sports. The interval between TTE and CMR was less than 24h in 93.3% of athletes; the interval between examinations was <30 days for all included athletes. Heart rate during TTE was higher than during CMR (57 bpm (51, 65) vs 53 bpm (48, 61), *p* < 0.001)

Athlete characteristics are presented in [**Table 1**]. TTE and CMR atrial strain and volumetric and function parameters are presented in [**Table 2**]. TTE measured larger minimal LA volumes (24 ml/m2 ±10 vs 22 ml/m2 ± 8, *p* = 0.014) and maximal LA volumes (63 ml/m2 ± 21 vs 52 ml/m2 ± 12, *p* = < .001) compared with CMR. Conversely, TTE measured a smaller minimal RA volume than CMR (29 ml/m2 ± 13 vs 37 ml/m2 ± 13, *p* = < .001). Max RA volume was comparable between TTE and CMR (63 ml/m2 ± 22 vs 62 ml/m2 ± 13, *p* = .328). TTE consistently measured less left- and right atrial myocardial deformation than CMR (LA reservoir strain: 34% ±6 vs. 44% ± 10, *p* < .001; LA conduit strain: 24% ± 5.4 vs. 31% ±8, *p* < .001; LA contractile strain: 9.7% ± 3.2 vs 13.3 ± 3.9, *p* < .001; RA reservoir strain: 32% ± 8 vs 53% ± 16, *p* < .001; RA conduit strain: 23% ±7 vs 37% ± 12, *p* < .001; RA contractile strain: 9.9% ± 3.4 vs 15.5% ± 5.8, *p* = < .001), respectively.

**Table 1.** Elite Athlete Characteristics.

| **Table 1 Elite Athlete Characteristics** |  |  |  |
| --- | --- | --- | --- |
| **Characteristic** | **Elite Athletes** |  | **P-value** |
|  | **Female**<br>**n = 98** | **Male**<br>**n = 96** |  |
| Age (years) | 30 (5) | 32 (8) | 0.2 |
| BSA (m2) | 1.79 (0.15) | 2.03 (0.18) | <0.001 |
| Sports Hours / Week (hours) | 18.9 (3.6) | 20.1 (4.6) | 0.13 |
| Professional Athlete years (years) | 13.4 (5.0) | 15.5 (8.2) | 0.13 |
| Sports Type (n, %) |  |  | 0.4 |
| Endurance | 36 (37%) | 45(47%) |  |
| Mixed | 43 (44%) | 39 (41%) |  |
| Power | 12 (12%) | 6 (6.3%) |  |
| Skill | 7 (7.1%) | 6 (6.3%) |  |
| Examination interval <24 hours (%) | 90,8% | 95,8% | 0.2 |
|  | **TTE**<br>**n = 194** | **CMR**<br>**n = 194** | **P-value** |
| LVEF (%) | 57.3 (4.9) | 56.1 (5.3) | 0.002 |
| LVEDV (ml) | 147 (34) | 225 (48) | <0.001 |
| LVEDVi (ml/m2) | 77 (15) | 117 (19) | <0.001 |
| LVESV (ml) | 63 (17) | 99 (26) | <0.001 |
| LVESVi (ml/m2) | 32 (7) | 51 (11) | <0.001 |
| LVSV (ml) | 85 (22) | 126 (28) | <0.001 |
| LVSVi (ml/m2) | 44 (10) | 66 (12) | <0.001 |
| BSA - Body Surface Area; LVEF - Left Ventricular Ejection Fraction; LVEDV - Left Ventricular End-Diastolic Volume; LVEDVi - BSA indexed LVEDV; LVESV - Left Ventricular End-Systolic Volume; LVESVi - BSA indexed LVESV; LVSV - Left Ventricular Stroke Volume; LVSVi - BSA indexed LVSV |  |  |  |

**Table 2.** Paired-Comparison of Bi-Atrial Volumes and Strain Between Transthoracic Echocardiography and Cardiac Magnetic Resonance Imaging.

| Characteristic | TTE<br>n = 194 | CMR<br>n = 194 | P-value |
| --- | --- | --- | --- |
| **Left Atrial** |  |  |  |
| Minimal LA Vol (ml/m<sup>2</sup>) | 24 (10) | 22 (8) | 0.014 |
| Maximal LA Vol (ml/m<sup>2</sup>) | 63 (21) | 52 (12) | <0.001 |
| Reservoir Strain (%) | 34 (6) | 44 (10) | <0.001 |
| Conduit Strain (%) | 24.0 (5.4) | 31 (8) | <0.001 |
| Contractile Strain (%) | 9.7 (3.2) | 13.3 (3.9) | <0.001 |
| **Right Atrial** |  |  |  |
| Minimal RA Vol (ml/m<sup>2</sup>) | 29 (13) | 37 (13) | <0.001 |
| Maximal RA Vol (ml/m<sup>2</sup>) | 63 (22) | 62 (17) | 0.328 |
| TTE RA Reservoir Strain (%) | 32 (8) | 53 (16) | <0.001 |
| TTE RA Conduit Strain (%) | 23 (7) | 37 (12) | <0.001 |
| TTE RA Contractile Strain (%) | 9.9 (3.4) | 15.5 (5.8) | <0.001 |

Female and male elite athlete characteristics for bi-atrial reservoir, conduit and contractile strain are shown in [**Supplemental Table 1**] and [**Supplemental Table 2**]. TTE specific results are shown in [**Supplemental Table 3**]. Across all components, CMR yielded higher strain values than TTE, indicating systematic underestimation by TTE for both LA and RA reservoir, conduit, and contractile function [**Supplemental Tables 1–2**].

### Agreement between STE and FT-CMR bi-atrial strain

We observed a weak correlation between STE and FT-CMR LA strain (LA reservoir strain: rho = .36, *p* < .001; LA conduit strain: rho = .36, *p* < .001; LA contractile strain: rho = .49, *p* < .001) [**Figure 2]**. We observed no correlation between STE and FT-CMR RA reservoir strain (rho = .06, *p* = .426) and RA conduit strain (rho = .13, *p* = .076), and a weak correlation for RA contractile strain (rho = .24, *p* < .001) [**Figure 3**].

**FIGURE 2.**
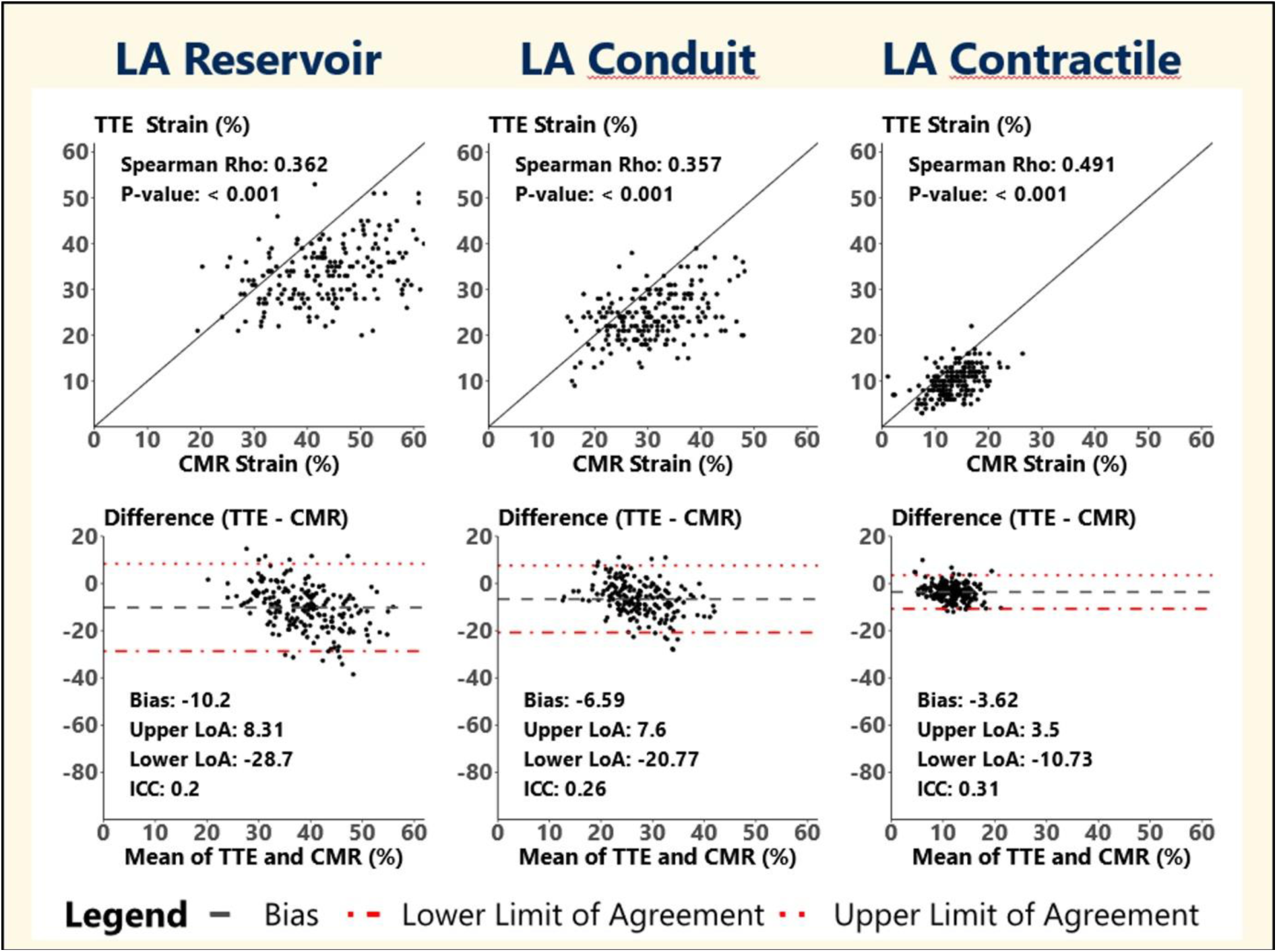
Correlation and Bland-Altmann Plots to Assess Agreement in Left Atrial Reservoir-, Conduit- and Contractile Strain for 2D Speckletracking Transthoracic Echocardiography (TTE) and Feature-Tracking Cardiac Magnetic Resonance Imaging (CMR)

**FIGURE 3.**
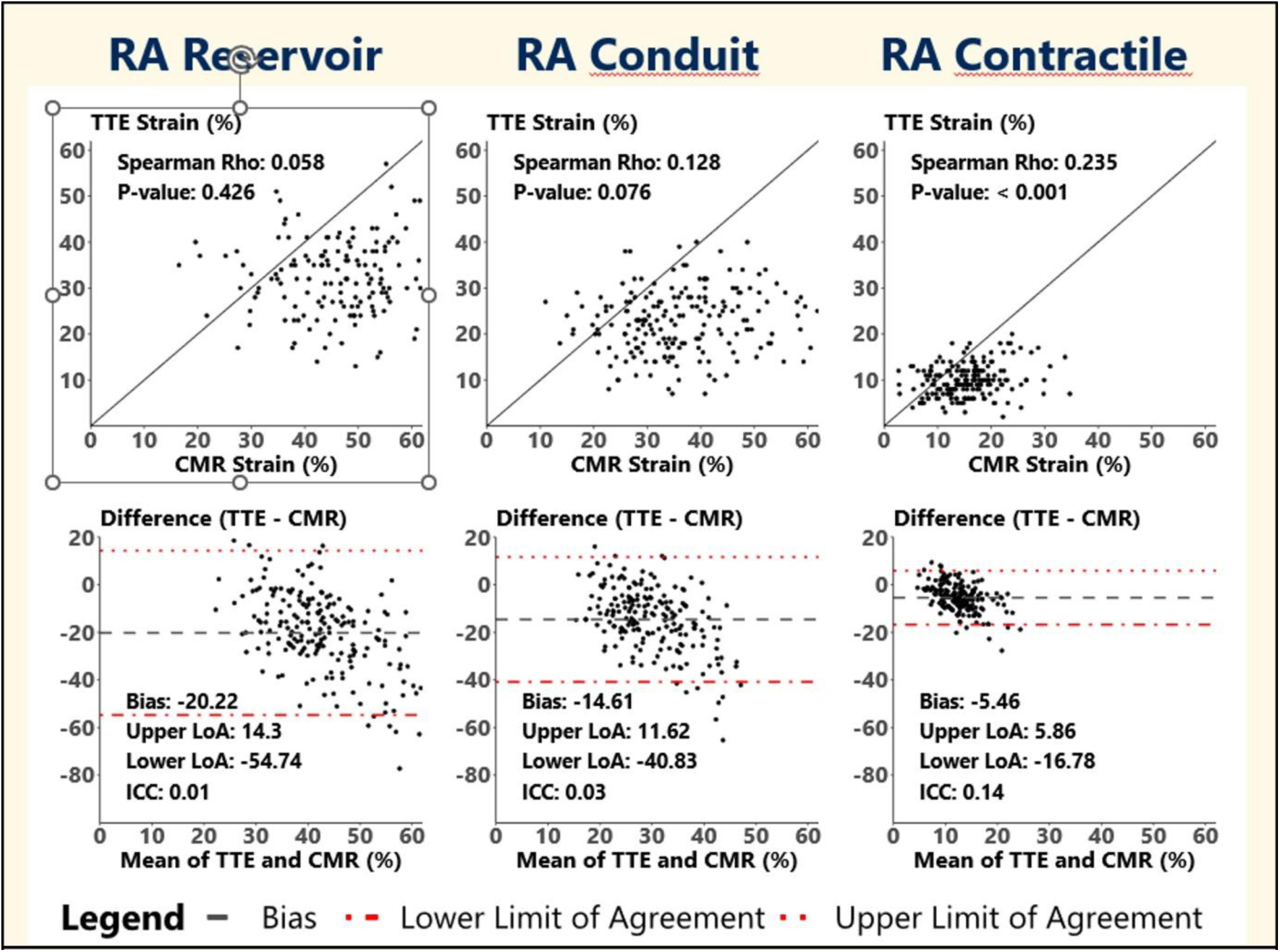
Correlation and Bland-Altmann Plots to Assess Agreement in Right Atrial Reservoir­ ’ Conduit- and Contractile Strain for 2D Speckletracking Transthoracic Echocardiography (TTE) and Feature-Tracking Cardiac Magnetic Resonance Imaging (CMR)

Left atrial strain demonstrated poor-to-moderate absolute agreement in two-way-mixed-effects model with single measure reliability (ICC[3,1]) (LA reservoir strain: 0.344 (95% CI: 0.213 – 0.460, *p* < .001; LA conduit strain: 0.39 (95% CI: 0.263 – 0.500, p < .001); LA contractile strain: 0.48 (95% CI: 0.359 – 0.580, p < .001) [**Figure 1**]. There was no agreement in RA reservoir strain (0.0178, 95% CI: -0.12 – 0.158, p = .4) and RA conduit strain (0.068, 95% CI: -0.073 – 0.21, p = .17. [**Figure 2].** There was poor agreement in RA contractile strain (0.218, 95% CI 0.080 – 0.48, *p* = 0.001). Remarkably, no correlation was observed between LA strain bias and LAVI (reservoir: *p* = 0.153; conduit: *p* = 0.070; contractile: *p* = 0.793) and RA strain bias and RAVI (reservoir: *p* = 0.286; conduit: *p* = 0.541; contractile: *p* = 0.217).

### Sex Differences in Atrial Strain

#### Left Atrial

On TTE, females demonstrated higher LA reservoir strain compared to males (41.5% ± 9.3 vs 37.1% ± 8.7, *p* < 0.01), and higher conduit strain (24.8% ± 6.9 vs. 22.3% ± 6.5, *p* = 0.03). On CMR, females showed higher values across all LA strain components (reservoir: 53.3% ± 9.0 vs 45.7% ± 8.6, *p* < 0.001; conduit: 32.3% ± 7.2 vs. 27.9% ± 7.0, *p* < 0.01; 20.7% ± 4.8 vs 18.2% ± 4.5, *p* = 0.01). For LA reservoir strain, bias was significantly greater in females than in males (−11.8% vs −8.6%, p = 0.016). Contractile strain bias was also significantly more negative in females (−4.4% vs −2.8%, p = 0.002). No significant sex difference was observed for conduit strain bias (−7.4% vs −5.7%, p = 0.099).

#### Right Atrium

On TTE, no significant sex differences were observed in RA strain between females and males (reservoir: 43.8% ± 9.4 vs 42.6% ± 9.1, *p* = 0.45; conduit: 24.2% ± 6.8 vs 23.9% ± 6.7, *p* = 0.72; contractile strain 19.6% ± 4.5 vs. 18.7% ± 4.3, *p* = 0.28). However, on CMR, females exhibited significantly higher RA reservoir 68.0% ± 9.5 vs 58.7% ± 10.2, *p* < 0.001) and contractile strain (25.7% ± 4.7 vs. 23.5% ± 5.0, *p* = 0.02), while conduit strain was similar (42.3% ± 6.7 vs. 40.6% ± 6.8, *p* = 0.11). RA strain bias was significantly greater in females for both reservoir (−24.3% vs −16.1%, p = 0.001) and conduit (−17.9% vs −11.2%, p < 0.001) components. No significant sex difference was observed for contractile strain bias (−6.4% vs −4.8%, p = 0.070). Overall, TTE underestimated RA strain to a greater extent in females, particularly for reservoir and conduit function.

### Comparison to Reference Values

Only a small percentage of athletes had STE and FT-CMR derived atrial strain values within the 95^th^ CI of the general population (LA reservoir: 2.6%; LA conduit: 8.3%; LA contractile strain: 0.5%). There was no difference in the rate of LA reservoir strain exceeding the reference limit of the general population between FT-CMR and STE (LA reservoir strain: 88.1% vs. 85.1%, χ² = 0.595, *p* = .440). However, FT-CMR demonstrated more LA conduit strain exceeding the reference limit than STE (LA conduit strain: 87.6% vs. 58.35%, χ² = 42.99, *p* < .001). Conversely, STE calculated LA contractile strain exceeded the reference limit more often than FT-CMR (88.14% vs. 95.9%, χ² = 6.759, *p* = .009).

### Interobserver ICC TTE

The interobserver agreement (ICC[2,1]) between the two raters for STE derived LA- and RA strain demonstrated good to excellent agreement for the LA (LA reservoir strain: 0.866, 95% CI 0.690–0.945; LA conduit strain: 0.875, 95% CI 0.711–0.949; LA contractile: 0.877, 95% CI 0.716–0.949). Similarly, RA strain also showed ICCs with good to excellent interobserver agreement (RA reservoir strain: 0.954, 95% CI 0.888–0.982; RA conduit strain: 0.947, 95% CI 0.871–0.979; RA contractile strain: 0.76, 95% CI 0.503–0.901).

## DISCUSSION

This study on TTE- and CMR-derived bi-atrial strain in elite athletes demonstrates substantial modality-dependent differences in bi-atrial size and function. The vast majority of athlete’s STE and FT-CMR derived strain values surpassed the reference values of the normal population, underlining the presence of considerable atrial remodeling in the athletic population. Although both imaging modalities yielded similar maximal RA volumes, CMR showed larger minimal RA volumes, while TTE demonstrated larger minimal and maximal LA volumes. Moreover, STE consistently yielded lower left- and right atrial strain compared to CMR. We observed poor-to-moderate agreement in LA strain between TTE and CMR. For the RA, we only observed poor agreement in contractile strain; there was no agreement in RA reservoir and conduit strain. Additionally, we demonstrated distinct sex-differences in bi-atrial strain in athletes. Female athletes showed higher LA strain across all components on both TTE and CMR, suggesting intrinsically higher LA strain in women. Remarkably, in contrast to the LA, the differences in RA strain between female- and male athletes were modality dependent. While we observed no differences in STE-derived RA strain between sexes, FT-CMR showed higher RA reservoir- and conduit strain in female athletes. These observations imply that CMR might be more sensitive to measure RA function. Additionally, strain bias was consistently greater in female athletes, particularly for RA reservoir and conduit strain, indicating that TTE might underestimate atrial strain in females, highlighting the importance of considering sex and imaging modality when assessing atrial function.

There are several potential explanations for the poor agreement in atrial strain measurements between STE and FT-CMR in athletes. These include differences in both image acquisition and quantification techniques, as well as anatomical and physiological factors such as breath-holding, body position, exercise, hydration status, and heart rate.

### Technical Differences in Atrial Volume Assessment

Although TTE usually underestimates true atrial volumes (23–26), our findings reveal a more complex pattern in athletes. Compared to CMR, TTE measures larger minimum and maximum LA volumes, but lower minimum RA volumes and comparable maximum RA volumes. CMR’s high-resolution imaging allows for more precise atrial volume assessment and personalized anatomical planning. In contrast, TTE is a two-dimensional modality that relies on acoustic windows and estimates atrial volumes using geometrical assumptions, which may not hold true in athletes with larger and more elongated atria, leading to an overestimation of atrial volumes (7).

### Strain Measurement Differences Between Modalities

Systematic intervendor- and intermodality dependent differences in LA strain have been reported (14). These may be caused by differences in myocardial motion tracking algorithms, as well as the regularization and presentation of data (14). In line with findings of Benjamin et al., we observed higher FT-CMR reservoir and conduit strain (17). However, while the study of Pathan et al. reported no difference in contractile strain between STE and FT-CMR (14), we found higher contractile strain values with FT-CMR. Additionally, our study demonstrated a proportional bias at higher FT-CMR strain values which is consistent with prior reports (14).

### Impact of Temporal Resolution

Part of the observed differences could be attributed to the higher temporal resolution of TTE (17). The superior frame rate of TTE allows more accurate capture of rapid changes in atrial size and function, particularly at higher heart rates (17). Conversely, the lower frame rate of CMR might underestimate atrial strain, especially in individuals with elevated heart rates, by missing the bimodal peaks of the atrial strain curve (14,17). However, as athletes generally have lower resting heart rates, this limitation is unlikely to explain the observed differences in our cohort.

### Anatomical Influences

Discrepancies in atrial volume assessment may arise from modality-specific differences in image acquisition and boundary detection. TTE frequently overestimates LA volume due to challenges in border definition in certain views, which can be affected by heterogeneous wall thickness, nonuniform segmental strain, and difficulties interpolating across pulmonary vein orifices and the LA appendage(27) (20). TTE is also more susceptible to artifacts and suboptimal acoustic windows, leading to foreshortening and inflated LA volume estimates. The RA poses even greater challenges due to its thin walls, complex geometry, and variable image quality, further hindering TTE’s ability to reliably track contours. Operator variability may further contribute to inconsistencies in atrial measurements between modalities.These issues are mitigated with CMR, where superior spatial resolution and well-defined borders enhance measurement precision and reproducibility (17).

### Breath-Holding and Body Position

Breath-holding during CMR image acquisition increases intrathoracic pressure, reducing venous return and affecting atrial filling, which may lead to lower RA and LA volume estimates (28). Additionally, CMR is performed in supine position while TTE is performed in left lateral decubitus position, altering atrial filling and deformation dynamics (28). Left lateral decubitus position promotes greater venous return due to reduced pressure on the inferior vena cava, leading to improved cardiac filling, yielding higher LA and RA reservoir strain values compared to the supine CMR position (28). The RA is especially pressure-sensitive and susceptible to body position and changes in venous return. As TTE is performed in left lateral decubitus position during normal breathing, this may cause fluctuations in preload and could explain why left atrial volume measurements can be greater with TTE.

### Physiological state

Although physiological factors like exercise and hydration status can influence strain, their impact on the observed variance in atrial strain is likely minimal, as 93.3% of athletes underwent TTE and CMRs within an hour of each other.

### Heart Rate

Athletes had a lower heart rate during CMR than during TTE, allowing for longer diastolic filling times and more complete atrial filling and emptying. Potentially, the prolonged diastolic filling contributed to the observed increase in reservoir, conduit, and to a lesser degree contractile strain, measured with FT-CMR, although the difference in heart rate was relatively small with a 5 bpm mean difference.

### Sex-specific Physiological Differences

We consistently observed higher LA strain in females across both modalities, which may reflect lower myocardial stiffness, less diffuse atrial fibrosis, and the favorable effects of estrogen on myocardial relaxation and compliance (29,30). Additionally, superior diastolic function in females may contribute to higher reservoir and conduit strain. In contrast, sex differences in RA strain were only detectable with CMR, suggesting that TTE may lack the sensitivity to capture subtle functional differences in the RA. The ability to detect sex-related differences may also be influenced by anatomical and physiological differences between sexes.

### Comparison to reference values

Over 90% of athletes had LA strain values above the 95^th^ % CI of the normal population, indicating that atrial remodeling is common in athletes. Importantly, in the context of athletes, exceeding standard reference values may reflect physiological remodeling rather than pathology. The proportion exceeding the reference limit for LA reservoir strain was similar between STE and FT-CMR, but FT-CMR more frequently detected elevated conduit strain, while STE more often detected elevated contractile strain. The FT-CMR reference values were derived from relatively small and potentially less diverse cohorts, causing narrower reference intervals which may not be generalizable to broader populations and may have affected these observations. These findings underscore the need for more robust and well-defined FT-CMR bi-atrial strain reference values, specifically in athletic populations.

### Limitations of Cross-Modality Comparison

The poor-to-moderate agreement in atrial volumes, left atrial (LA) strain, and the lack of agreement in right atrial (RA) strain between TTE and CMR underscores the need for caution when interpreting atrial strain across imaging modalities. Discrepancies in atrial volume assessment and border tracking, along with technical, methodological, anatomical, and physiological differences, limit the comparability of atrial function metrics between STE and FT-CMR. Our findings demonstrate that TTE and CMR cannot be used interchangeably to evaluate atrial function in athletes, and that measurements obtained from one modality cannot be extrapolated to the other. This underscores the need to interpret bi-atrial strain values using modality- and sex specific reference standards tailored to the athletic population.

### Future

Future studies should focus on establishing robust, sport-specific, and modality-specific reference values for atrial strain in athletic populations. At present, it remains unclear whether TTE or CMR strain offers greater prognostic value. Future studies should aim to establish associations between atrial strain and clinical outcomes (e.g., VO₂ max) to address this question. This is particularly important to refine pre-participation screening, guide longitudinal follow-up, and support tailored, sport-specific clinical decision-making regarding eligibility and risk stratification in athletes.

## CLINICAL IMPLICATIONS

Considering the substantial modality-dependent differences in atrial strain, results from STE studies in elite athletes cannot be extrapolated tot CMR or vice versa. Clinicians should be cautious when comparing atrial strain values across modalities as TTE systematically measured lower bi-atrial strain values. The poor-to-moderate agreement between STE and FT-CMR highlights the necessity of modality specific reference values in elite athletes. When assessing atrial function over time, maintaining consistency in the imaging modality is important to ensure reliable trend analyses, particularly in populations prone to cardiac remodeling or at risk for adverse cardiac effects.

## STRENGTHS AND LIMITATIONS

There are several strengths to our study. First, we included a substantial number of elite athletes from different athletic backgrounds, increasing generalizability beyond the scope of endurance athletes. Second, we included equal groups of female and male athletes, ensuring that our findings are also generalisable to female athletes. Lastly, the TTE and CMR were both performed within a short time frame, limiting the influence of hydration status and other physiological factors.

Some aspects of our study warrant consideration. First, the bi-atrial strain values were derived from a single measurement rather than multiple acquisitions. While this may introduce some variability, it better reflects routine clinical practice where single measurements are typically used for decision making. Second, prior studies have demonstrated that atrial strain measurements are susceptible to interobserver variability, influencing the consistency and reproducibility of results. In our cohort STE-derived atrial strain was assessed by two independent observers but interobserver ICC analysis showed good-to-excellent agreement between the two observers. Lastly, FT-CMR derived strain was assessed by a single observer, further limiting the influence of interobserver variation.

## CONCLUSION

Our study demonstrates significant modality-dependent differences in bi-atrial strain in elite athletes, with poor to moderate agreement between longitudinal STE and FT-CMR left atrial strain, and a lack of agreement in right atrial strain. STE consistently yielded lower strain values, suggesting that each modality captures different aspects of atrial function. Given these discrepancies, STE-derived values cannot be extrapolated to FT-CMR, underscoring the limited interchangeability of TTE and CMR for atrial strain assessment. As bi-atrial strain is highly modality-dependent, strain values should always be interpreted in the context of modality-specific reference values to ensure accurate evaluation of atrial function in athletes.

## Data Availability

Data are available upon reasonable request

## SUPPLEMENTAL MATERIAL

**Supplemental Table 1.** Female Elite Athlete Reference Ranges for Speckle Tracking Echocardiography (STE) and Feature Tracking Cardiac Magnetic Resonance Imaging (FT-CMR) Derived Bi-Atrial Strain.

| Reference Range |  |  |
| --- | --- | --- |
| | Mean ± SD | Percentile (2,5-97,5%) |
| **Left Atrial Strain** |  |  |
| **STE:** |  |  |
| Reservoir | 35.4 ± 6.7 | (22.3 – 50.2) |
| Conduit | 25.6 ± 5.5 | (15.0 – 36.6) |
| Contractile | 9.8 ± 2.9 | (5.0 – 16.0) |
| **FT-CMR:** |  |  |
| Reservoir | 47.2 ± 9.1 | (30.3 – 63.7) |
| Conduit | 33.0 ± 7.1 | (19.4 – 47.8) |
| Contractile | 14.2 ± 3.9 | (7.7 – 21.4) |
| **Right Atrial Strain** |  |  |
| **STE:** |  |  |
| Reservoir | 32.7 ± 8.2 | (15.4 – 47.7) |
| Conduit | 22.9 ± 6.9 | (10.4 – 36.7) |
| Contractile | 9.7 ± 3.2 | (4.4 – 17.0) |
| **FT-CMR:** |  |  |
| Reservoir | 56.9 ± 14.6 | (29.9 – 86.0) |
| Conduit | 40.9 ± 11.3 | (20.5 – 62.9) |
| Contractile | 16.1 ± 5.4 | (7.0 – 28.3) |

**Supplemental Table 2.** Male Elite Athlete Reference Ranges for Speckle Tracking Echocardiography (STE) and Feature Tracking Cardiac Magnetic Resonance Imaging (FT-CMR) Derived Bi-Atrial Strain.

| **Reference Range** |  |  |
| --- | --- | --- |
| | Mean ± SD | Percentile (2,5-97,5%) |
| **Left Atrial Strain** |  |  |
| **STE:** |  |  |
| Reservoir | 32.0 ± 5.8 | (22.0 – 42.9) |
| Conduit | 22.4 ± 4.8 | (13.0 – 31.9) |
| Contractile | 9.6 ± 3.5 | (4.0 – 16.6) |
| **FT-CMR:** |  |  |
| Reservoir | 40.6 ± 9.2 | (24.4 – 58.4) |
| Conduit | 28.2 ± 7.2 | (15.9 – 43.5) |
| Contractile | 12.4 ± 3.7 | (5.7 – 18.3) |
| **Right Atrial Strain** |  |  |
| **STE:** |  |  |
| Reservoir | 32.2 ± 8.6 | (17.0 – 49.0) |
| Conduit | 22.2 ± 7.0 | (10.0 – 36.9) |
| Contractile | 10.1 ± 3.7 | (4.4 – 18.0) |
| **FT-CMR:** |  |  |
| Reservoir | 48.3 ± 15.7 | (23.0 – 84.7) |
| Conduit | 33.4 ± 11.6 | (16.1 – 64.9) |
| Contractile | 14.9 ± 6.1 | (3.8 – 28.2) |

**Supplemental Table 3.** Transthoracic Echocardiography Results.

|  | Elite Athletes |  | P-value |
| --- | --- | --- | --- |
|  | Female<br>n = 98 | Male<br>n = 96 |  |
| LV Ejection Fraction (%) | 57.6 (4.4) | 57.1 (5.3) | >0.9 |
| LV Internal Diameter (Diastole, mm) | 5.15 (0.47) | 5.65 (0.44) | <0.001 |
| LV Internal Diameter (Systole, mm) | 3.34 (0.51) | 3.67 (0.74) | 0.046 |
| LV Posterior Wall Thickness (mm) | 0.88 (0.16) | 0.99 (0.17) | <0.001 |
| Relative Wall Thickness | 0.36 (0.23) | 0.76 (3.72) | 0.044 |
| Tricuspid Annular Plane Systolic<br>Excursion (TAPSE) (mm) | 2.73 (0.41) | 2.85 (0.51) | 0.2 |
| E-Wave Velocity (m/s) | 0.88 (0.17) | 0.80 (0.17) | <0.001 |
| A-Wave Velocity (m/s) | 0.44 (0.11) | 0.39 (0.10) | 0.004 |
| E/A Ratio | 2.09 (0.58) | 2.16 (0.63) | 0.3 |
| E/e' Ratio | 5.60 (1.04) | 5.03 (1.13) | <0.001 |
| Left Atrial Area (cm<sup>2</sup>) | 18.9 (3.3) | 21.7 (4.1) | <0.001 |
| Left Atrial Maximal Volume (mL) | 20 (8) | 29 (11) | <0.001 |
| Left Atrial Minimal Volume (mL) | 20 (8) | 29 (11) | <0.001 |
| Tricuspid Regurgitation Max PG<br>(mmHg) | 18.2 (4.3) | 21.3 (5.4) | 0.001 |

